# Efficacy and safety of colchicine for the treatment of osteoarthritis: protocol for a systematic review and meta-analysis of intervention trials

**DOI:** 10.1101/2020.11.20.20226589

**Authors:** Pablo Molina-Garcia, Ambrish Singh, Salman Hussain, Sidharth Das, Benny Antony

## Abstract

Colchicine, in the form of *Colchicum autumnale*, has been used to treat joint swelling for centuries. The anti-inflammatory action o colchicine and the higher prevalence of calcium pyrophosphate crystal deposition (CPPD) in osteoarthritis (OA) joints have led to th use of colchicine as a potential treatment for OA, however, the quality of the evidence on this regards is still limited. The curren protocol aims to conduct a systematic review and meta-analysis proving the efficacy and safety of colchicine for the treatment of adul patients with OA. For that purpose, Scopus®, Web of Sciences®, Medline® and Cochrane Library® will be inspected for the availabl trials testing the efficacy and safety of colchicine for the treatment of any type of OA (e.g., hand, knee or hip). Cochrane tool will b used to assess the risk of bias of included trials. A meta-analysis of dichotomous (e.g., adverse events) or continuous data (e.g., mea difference in pain scale) will be performed depending on the data reporting. Review Manager 5 (RevMan) and RStudio Version 1.2.133 will be used to conduct the statistical analysis.

## 1. Background and rationale

Osteoarthritis (OA) incidence continues to increase and is expected to be one of the most prevalent diseases in developed countries in the coming decades.^1^ Severe pain and loss of physical function are common symptoms associated with OA and are among the leading cause of disability and time off work worldwide.^2^ To date, there are no available disease-modifying treatments to revert OA, and the usual pharmacological management is limited to non-steroidal anti-inflammatory drugs (NSAIDs) that palliate its symptoms.^3^ These drugs, however, have demonstrated modest benefits on pain and functional outcomes and might lead to adverse effects in elders and people with comorbidities.^3^ All this, reinforce the need to investigate further pharmacological treatments that are both effective and safe against OA.

Colchicine is a medication extracted from the *Colchicum autummale* plant that has been used to treat ailments related to joint swelling for thousands of years. Nowadays, it is one of the main treatment options against some rheumatic diseases such as gout and familial Mediterranean fever.^4^ Its anti-inflammatory effects seems promising in the treatment of OA. A systematic review and meta-analysis revealed the positive effects of colchicine in pain reduction and physical functioning of adult patients with knee OA.^5^ Nonetheless, the quality of this evidence is limited since the review only included five randomized control trials with small sample size and considerable risk of bias. Given that this systematic review of the literature was conducted in November 2016, an update on this topic is justified to improve the quality of the evidence about the efficacy and safety of colchicine on any type of OA.

## 2. Objective

To evaluate the efficacy and safety of colchicine on pain, physical function, and adverse consequences in patients with OA.

## 3. Eligibility criteria

PICOS strategy will be used to determine the eligibility of studies based on: Population, Interventions, Comparators, Outcomes, and Study design.

### Population (P)

Adult participants older than 18 years, of any sex, and diagnosed with OA according to the American College of Rheumatology criteria or similar approaches.^6^

### Interventions/exposure (I)

Oral alone or in combination with other conventional drugs such as Paracetamol or NSAIDs.

### Comparator/control (C)

Active control or placebo control group.

### Outcomes (O)

Pain, physical function, imaging biomarkers (x-ray and/or MRI structural measures), biochemical markers, medication change, quality of life (QoL), and adverse events.

### Study design (S)

Randomised control trials, quasi-randomised, and non-randomised control trials with blinded or non-blinded design.

## 4. Data sources

Scopus®, Web of Science®, Medline®, and Cochrane Library® databases will be searched for this systematic review. The abstracts of last two year’s key conferences in the field of OA and rheumatology [e.g., European League Against Rheumatism (EULAR), Osteoarthritis Research Society International (OARSI), American Academy of Orthopaedic Surgeons (AAOS), and American College of Rheumatology (ACR)], as well as the reference list of included studies, and related systematic reviews and meta-analysis will be hand searched.

## 5. Search strategy

Scopus®, Web of Science®, Medline®, and Cochrane Library® databases will be searched using the following representative search strategy.

|  |  |
| --- | --- |
| Scopus® | **Population**<br>TITLE-ABS-KEY ( ( osteoarthr* OR "arthrosis" OR "artrosis" ) ) |
|  | **Intervention**<br>TITLE-ABS-KEY ( ( "colchicine" OR "colchicina" ) ) |
| Web of Sciences®<br>(All databases) | **Population**<br>TS=(osteoarthr* OR "arthrosis" OR "artrosis") |
|  | **Intervention**<br>TS=("colchicine" OR "colchicina" ) |
| Medline® | **Population**<br>All Fields (osteoarthr* OR "arthrosis" OR "artrosis") |
|  | **Intervention**<br>All Fields ("colchicine" OR "colchicina" ) |
| Cochrane Library® | **Population**<br>All Text (osteoarthr* OR "arthrosis" OR "artrosis") |
|  | **Intervention**<br>All Text ("colchicine" OR "colchicina" ) |

## 6. Study selection and data extraction

Two reviewers will individually perform the study selection process, and any discrepancy will be resolved through discussions and consensus. The systematic review software “Covidence” (Veritas Health Innovation) will be used in order to facilitate the study selection process.

Relevant data of included studies will be extracted by one researcher and double-checked by a second reviewer. Information will be extracted as follow: 1) author’s name and year of publication, 2) study design 3) sample size and population characteristics, 4) method used to identify/grade OA, 5) colchicine treatment (i.e., dose, administration manner, frequency, duration and follow up) 6) description of placebo/control, 7) outcomes included (e.g., pain, physical functioning, and QoL), 8) main findings.

## 7. Risk of bias

The risk of bias of included studies will be individually tested by two researchers, according to the *Cochrane Handbook for Intervention Studies* guidelines. The criteria applied will be the following: 1) random sequence generation, 2) allocation concealment, 3) blinding of participants and personnel, 4) blinding of outcome assessment, 5) incomplete outcome data, 6) selection reporting, 7) attention, 8) compliance and 9) other potential sources of bias. Review Manager software (v. 5.4) will be used to guide the risk of bias assessment, and any disagreement will be resolved by the discussion with senior authors.

## 8. Statistical analysis

A meta-analysis of dichotomous (e.g., pain reduction or physical function improvement) or continuous data (e.g., mean difference in pain scale or physical function score) will be performed depending on how most studies report the data. In the case of a meta-analysis of dichotomous data, results will be summarized by means of the pooled relative risks (RR) estimate (and 95% CI interval) and number needed to treat (NNT),^7^ while for the meta-analysis of continuous data, the standard mean differences (i.e., Hedges’ g or Cohen’s d effect sizes depending on the sample size) will be used. Heterogeneity will be assessed with the Higgins I^2^ statistic and p values, being classified as not important (0-40%), moderate (30-50%), substantial (50-75%), or considerable (75-100%).^8^ In the presence of substantial or considerable heterogeneity (I^2^ > 50%), the random-effects model will be used, while if not important or moderate heterogeneity is found (I^2^ < 50%), the fixed-effects model will be used.^9^ Sensitivity analyses will be performed by removing studies one at a time and testing whether the overall effect size (e.g., z-value) is significantly modified in magnitude or direction. This approach will provide information about the robustness of the synthesis of the evidence. A funnel plot and the Egger regression asymmetry test (a level of <0.10) will be used to assess publication bias.^10^ In the case that ten or more studies will be included, meta-regression analysis will be performed to test how the study characteristics (e.g., sex and age of participants, colchicine dose or treatment duration) are associated with the colchicine treatment effects. Analyses will be performed using the Review Manager Version 5.4 (The Nordic Cochrane Center, The Cochrane Collaboration, 2014, Copenhagen, Denmark) and RStudio Version 1.2.1335 (PBC, Vienna, Austria) with the metaphor and meta (version 4.9-5) packages.

## Data Availability

This preprint is a protocol for a systematic review and meta-analisis and there are no current data available.

## Notes

### Competing Interest Statement

The authors have declared no competing interest.

### Clinical Trial

NA

### Funding Statement

No external funding was received

